# Cardiac Rehabilitation and Functional Capacity Improvement Montana Outcomes Project Cardiac Rehabilitation Registry Findings

**DOI:** 10.64898/2026.04.20.26351126

**Authors:** Leah Claus, Michael McNamara, Carrie Oser, Crystelle Fogle, Brenda Canine

## Abstract

Cardiovascular disease (CVD) remains the leading cause of mortality in the United States, despite being largely preventable through effective management of risk factors. This study evaluates the impact of Phase II cardiac rehabilitation (CR) on functional capacity and quality of life, using data from the Montana Outcomes Project Cardiac Rehabilitation Registry. Functional capacity improvements were assessed via the six-minute walk test (6MWT) and Dartmouth COOP questionnaire, with statistical analyses exploring the influence of CR session attendance, demographic factors, and referring diagnoses. Results demonstrated significant gains in 6MWT, with a mean improvement of 330.73 feet (p < .0001), and quality of life scores across all subgroups. A dose-response relationship was observed, indicating greater improvements with increased CR sessions (p < .0001), though diminishing returns were observed beyond 24-35 visits. Demographic factors and complex conditions influenced outcomes, underscoring the need for tailored strategies to enhance CR access and effectiveness. These findings highlight the critical role of CR in improving patient outcomes and emphasize the importance of addressing barriers to participation in underserved populations.

## Introduction

Cardiovascular disease (CVD) remains the leading cause of mortality in the United States since 1921^1–3^. Yet, its etiology suggests that CVD is largely preventable, as modified risk factors account for over 90% of the risk for developing the CVD^4^. Prioritizing healthy lifestyle behaviors, such as regular physical activity, smoking cessation, and dietary modifications, has the potential to significantly reduce the burden of CVD.

Cardiac rehabilitation (CR) serves as a highly effective secondary prevention model for managing CVD and mitigating its associated risk factors^5^. Phase II CR programs offer a comprehensive approach that combines structured exercise with education and behavioral counseling. These programs address critical health outcomes, including improvement in functional capacity, blood pressure regulation, tobacco cessation, and mental health through depression screenings. Importantly, regular participation in CR is associated with a 32% reduction in all-cause mortality^6^.

One key performance metric of CR programs is the improvement in functional capacity, often measured by the six-minute walk test (6MWT). Despite the documented benefits of CR, knowledge gaps persist regarding how specific factors – such as the number of sessions completed, demographic characteristics, and referring diagnoses – may influence these outcomes.

This study seeks to address these gaps by investigating the impact of phase II CR on functional capacity. The specific aims of this research are threefold: (1) to quantify the average improvement in functional capacity among CR participants, (2) to evaluate whether a dose-response relationship exists between the number of CR visits and functional capacity improvements, and (3) to examine whether demographic factors or referring diagnoses affect these outcomes. By addressing these objectives, this study provides valuable insights into optimizing the delivery and effectiveness of CR programs while promoting equitable access to care.

## Methods

### CR REGISTRY AND PATIENT DATA COLLECTION

These data were collected from the Montana Outcomes Project, a data registry developed and coordinated by the Cardiovascular Health Program within the Montana Department of Public Health and Human Services. The registry encompasses over 100 submitting programs across the United States. A data use agreement was formalized with Touro College of Osteopathic Medicine. This project was determined to be non-human subjects research following the Touro University New York IRB Human Subjects Research Determination form. The project aims to deliver a comprehensive outcomes program that can provide feedback and enable programs to compare their individual data with a collective group. This drives quality improvement initiatives to ultimately enhance patient care.

### FUNCTIONAL CAPACITY COLLECTION

Six-minute walk test (6MWT) data were used as a reliable method to assess functional capacity improvement in the phase II CR population^7^. The Dartmouth COOP (COOP) questionnaire was used to assess functional health and quality of life. The self-questionnaire includes 9 questions, each with a score of 1-5, to total 9 to 45 points. Lower scores represent greater functional capacities and quality of life. The reliability and validity of the COOP have been previously demonstrated^8,9^.

### PATIENT DEMOGRAPHICS

Patient demographic data were recorded for sex, race (White, American Indian, Black, Asian, and Other), Hispanic ethnicity, number of phase II visits completed, presence of insurance and, referring diagnoses of diabetes and CVD. Patients completing 12-63 phase II sessions were included and were stratified by completion of 12-23, 24-35, 36+ sessions. 12 sessions were used as the lower limit for inclusion in the current study. With respect to disease classifications, patients were classified based on the primary referring diagnoses.

### STATISTICAL ANALYSES

The data were analyzed using R (R Core Team, 2024). Data are reported as mean ± SD. Pre- and post-data for 6MWT and COOP for the group, sex, race, ethnicity, referring diagnoses, and presence of insurance were examined using analysis of variance. Groups with equal variances were assessed using two sample-t-tests. Pairwise comparisons were made using Wilcoxon rank sum test. Multiple regression analysis was used to identify any relationships with multiple confounding variables. The level of significance was set at p < 0.05.

## Results

According to the inclusion/exclusion methodology, 4,147 patients completing 12-63 CR sessions, pre- and post-6MWT, and pre- and post-Dartmouth COOP are reported. Patient characteristic data are presented in Table 1. Patients averaged 68.74 yr and completed 29.57 CR sessions. Males (n = 2,935) represented approximately 70% of the patient population, and 93.6% of patients were White (n = 3,907). Patients were most often referred after revascularization and/or repair procedures (coronary artery bypass graft [CABG] = 830, percutaneous coronary intervention [PCI] = 997), myocardial infarction [MI]/CABG = 161, MI/PCI = 757. The dataset included 1,187 individuals with diabetes. Most participants did not use tobacco (84.3%) and had medical insurance (97.3%).

**Table 1.** Participant Characteristics (n=4,147)^a^.

|  |  |
| --- | --- |
| *Phase II visits, d* | 29.57 ± 8.71 |
| *Age, yr* | 68.74 ± 11.20 |
| *Sex* |  |
| *Female* | 1,212 (29.2) |
| *Male* | 2,935 (70.8) |
| *Race* |  |
| *White* | 3,807 (91.8) |
| *American Indian* | 54 (1.3) |
| *Black* | 49 (1.2) |
| *Asian* | 27 (0.7) |
| *Other* | 61 (1.5) |
| *Ethnicity* |  |
| *Hispanic* | 88 (2.1) |
| *Referring diagnoses* |  |
| *MI* | 264 (6.4) |
| *MI/CABG* | 161 (3.9) |
| *MI/PCI* | 757 (18.3) |
| *CABG* | 830 (20.0) |
| *PCI* | 997 (24.0) |
| *Angina* | 118 (2.8) |
| *Heart Failure* | 620 (15.0) |
| *Systolic Type* | 352 (56.8) |
| *Diastolic/right side* | 183 (29.5) |
| *Valve replacement/repair* | 524 (12.6) |
| *Transplant* | 19 (0.5) |
| *LVAD* | 14 (0.3) |
| *PAD* | 42 (1.0) |
| *TAVR* | 320 (7.7) |
| *Other* | 113 (2.7) |
| *Diabetic status* |  |
| *Diabetic* | 1,187 (28.6) |
| *Nondiabetic* | 2,960 (71.4) |
| *Tobacco status* |  |
| *Current Tobacco User* | 243 (5.9) |
| *Former Tobacco User* | 204 (4.9) |
| *Non-Tobacco User* | 3,497 (84.3) |
| *Insurance status* |  |
| *Has insurance* | 4,033 (97.3) |
| *No insurance* | 8 (0.2) |

Abbreviations: CABG, coronary artery bypass graft; LVAD, left ventricular assist device; MI, myocardial infarction; PAD, peripheral artery disease; PCI, percutaneous coronary intervention; TAVR, transthoracic aortic valve replacement.
<sup>a</sup>Data are presented as mean ± SD or n (%).

### SEX, RACE, AND ETHNICITY

6MWT values pre- and post-CR number of phase II visits, sex, race, and ethnicity distribution are presented in Table 2. Results demonstrated significant gains in 6MWT, with a mean improvement of 330.73 feet (p < .0001). Additionally, there were significant increases in the 6MWT score after phase II CR for all variables (p < .0001). Stratification for the number of phase II CR visits (12-23, 24-35, 36+) revealed that 6MWT values were improved in all tiers (p < .0001). Further pairwise comparison revealed significant difference between group means of 6MWT percent change for 12-23 and 24-35 phase II CR visits (p < .0001). The same was true when comparing the 12-23 to 36+ group (p < .0001). There was no difference in group means between 24-35 and 36+ groups (p = 0.26). These are reflected in Figure 1.

**Table 2.**
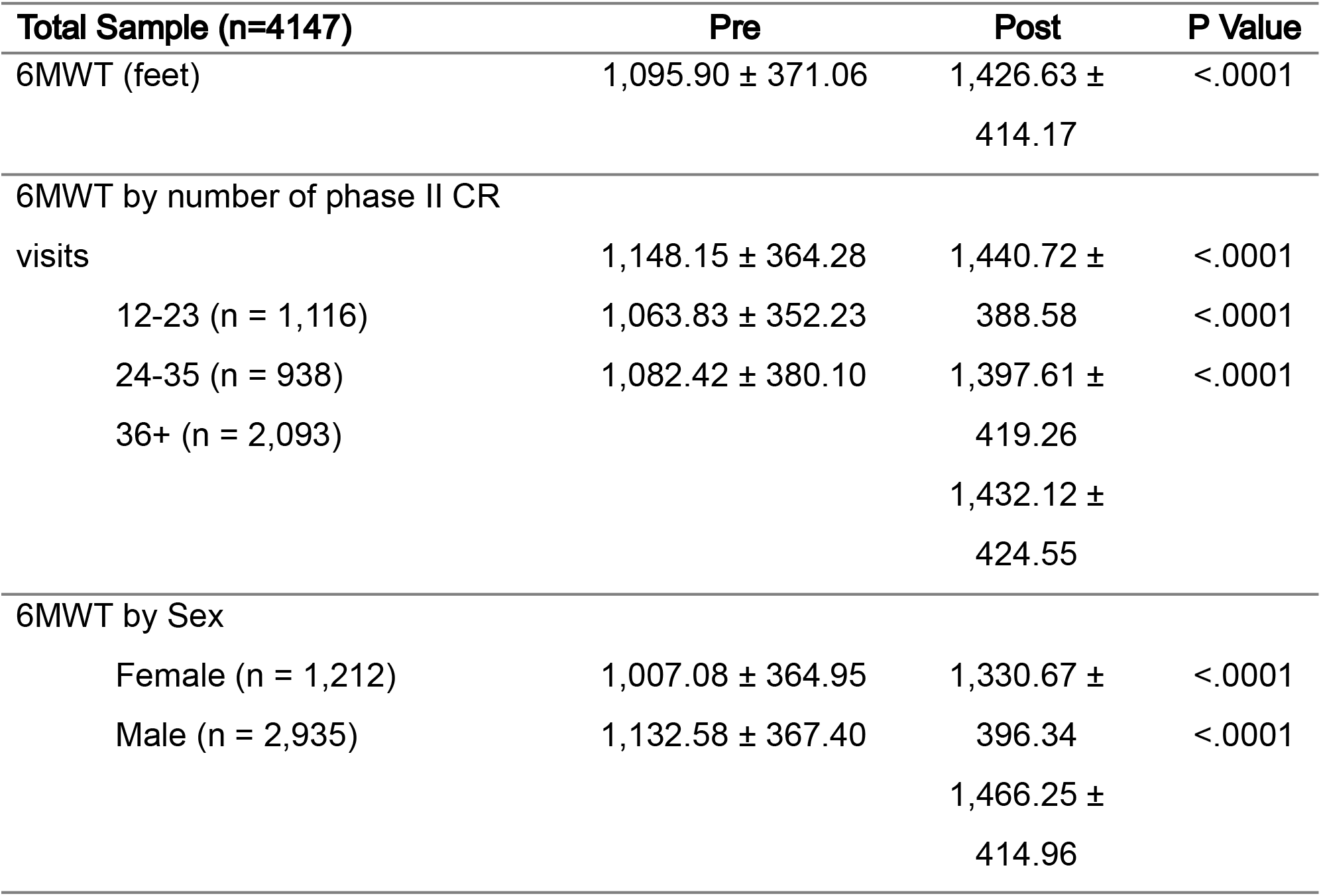
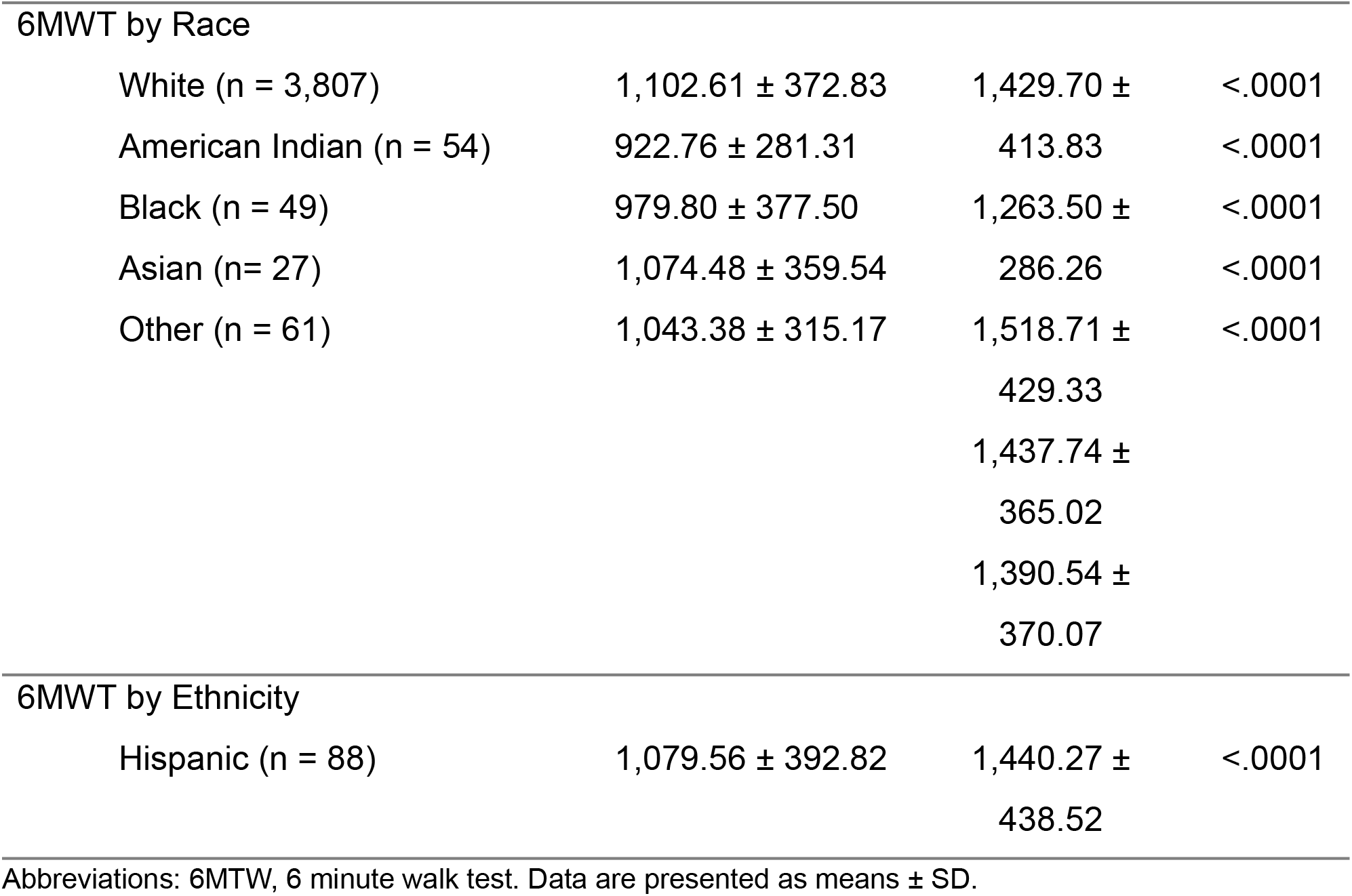
Six-Minute Walk Test (6MWT) Values Pre and Post 12-63 Sessions of Cardiac Rehabilitation for Total Group, Sex, Race, and Ethnic Distributions.

**Figure 1.**
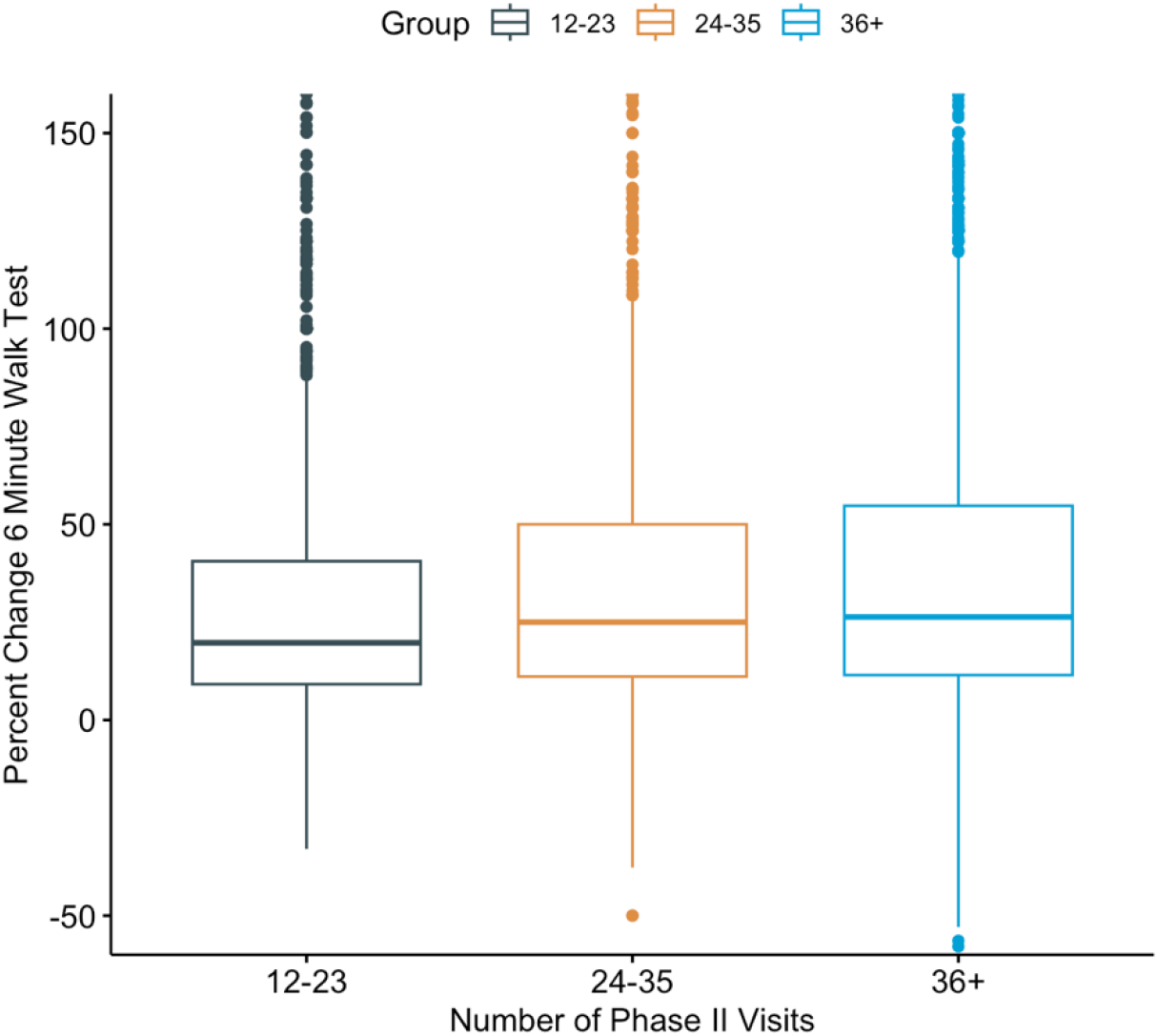
Percent Change for 6MWT Across Groups for Phase II CR Visits

The 6MWT mean percent change for Black participants was significantly different than the group means for all other races (p<.01). When examining racial differences, Black patients had a lower starting value for pre-phase II CR visits, however this was not statistically significant. There were no differences in group means for ethnicity.

Paired t-tests illustrated significant differences (p < .05) in pre and post Dartmouth scores across all variables (phase II CR visits, Sex, Race, Ethnicity, etc.) (Table 3). There were no notable trends illustrating a change in Dartmouth COOP score with the number of phase II CR visits completed.

**Table 3.**
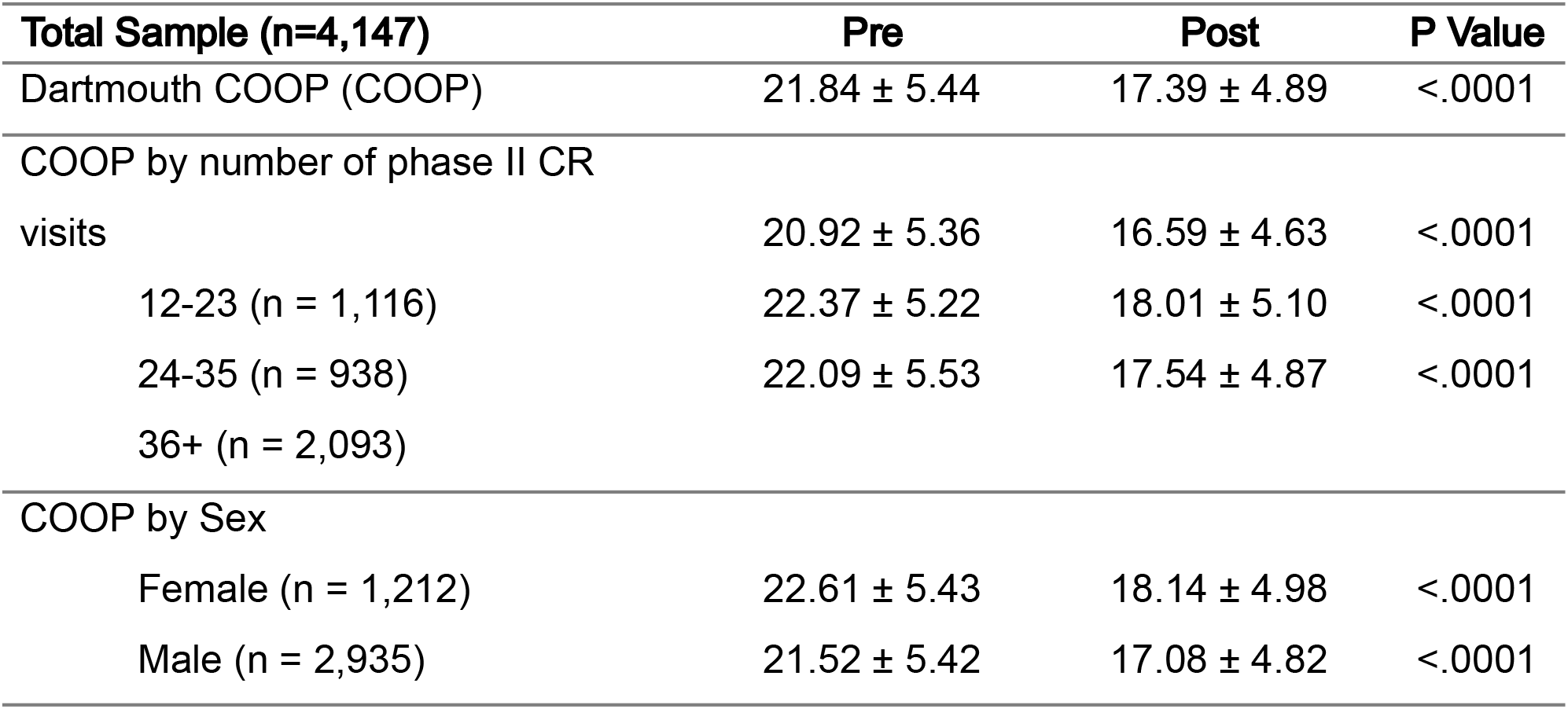
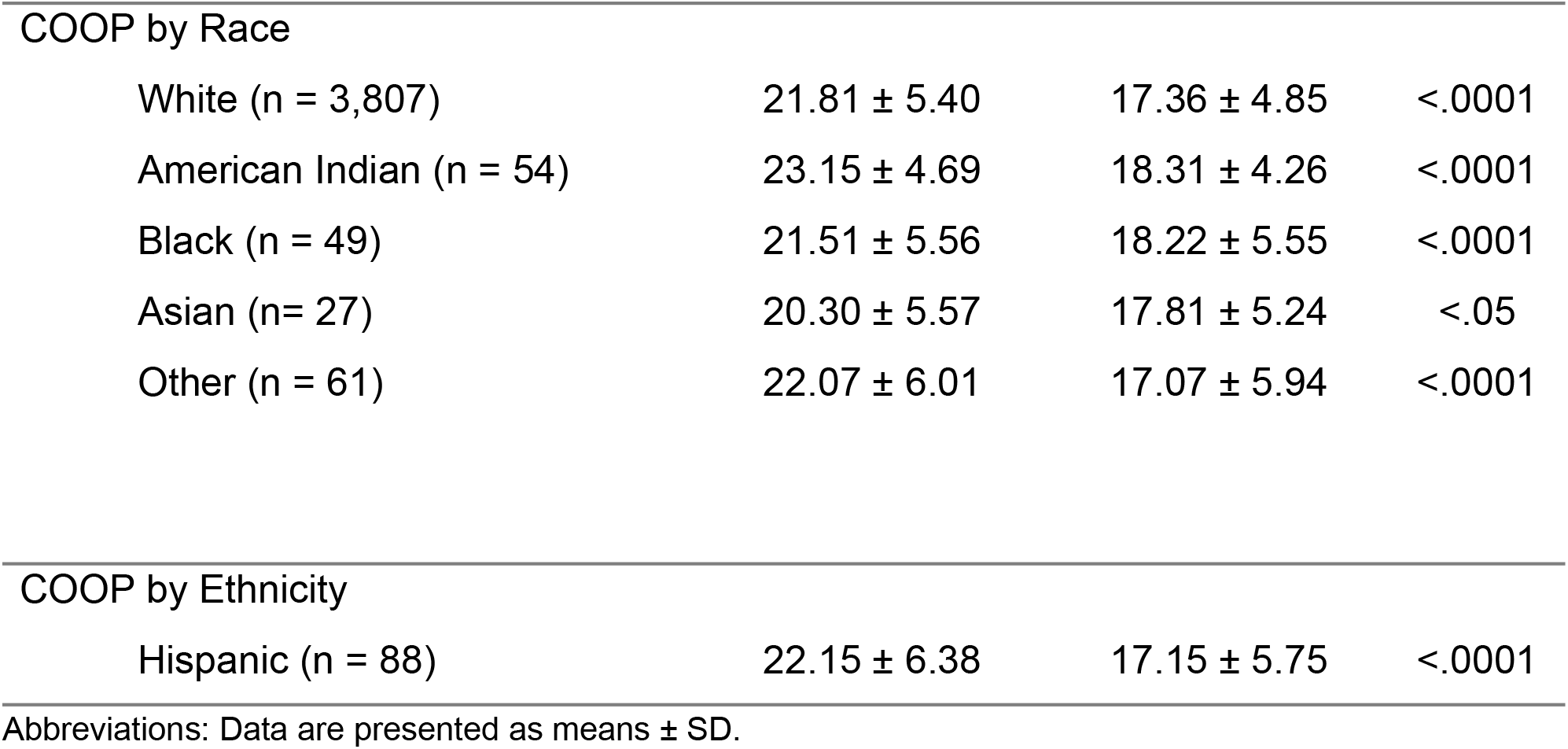
Dartmouth COOP Pre and Post 12-63 Sessions of Cardiac Rehabilitation for Total Group, Sex, Race, and Ethnic Distributions.

### CARDIOVASCULAR AND DIABETIC DIAGNOSIS

6MWT Values pre- and post-CR for the primary referring cardiac and diabetes diagnoses are reported in Table 4. 6MWT scores improved across most referring and diabetes diagnoses (p < .0001). Patients with LVAD as their referring diagnosis also significantly improved their 6MWT scores, but at p < .01. Patients with diagnoses for transplant, left ventricular assist device (LVAD), and peripheral artery disease (PAD) had fewer submitting participants compared to other groups. There was no difference in 6MWT scores for patients with diabetes compared to those without diabetes (p = .08).

**Table 4.** 6 Minute Walk Test (6MWT) Values Pre and Post 12-63 Sessions of Cardiac Rehabilitation for the Referring Diagnosis and Diabetes.

| Referring Diagnosis | Pre | Post | P Value |
| --- | --- | --- | --- |
| MI (n = 264) | 1,148.72 ± 370.57 | 1,440.27 ± 412.54 | <.0001 |
| MI/CABG (n = 161) | 1,002.88 ± 359.67 | 1,375.04 ± 348.38 | <.0001 |
| MI/PCI (n = 757) | 1,156.44 ± 370.47 | 1,498.36 ± 418.35 | <.0001 |
| CABG (n = 830) | 1,046.09 ± 338.57 | 1,425.62 ±<br>378.41 | <.0001 |
| PCI (n= 997) | 1,156.95 ± 388.50 | 1,447.09 ±<br>423.08 | <.0001 |
| Angina (n= 118) | 1,111.49 ± 345.82 | 1,325.73 ±<br>372.83 | <.0001 |
| Heart Failure (n = 620) | 1,027.79 ± 357.99 | 1,357.40 ±<br>406.28 | <.0001 |
| Systolic heart failure (n = 352) | 1,023.64 ± 357.22 | 1,360.15 ±<br>402.96 | <.0001 |
| Diastolic/right heart failure (n = 183) | 1,036.00 ± 369.77 | 1,396.50 ±<br>421.56 | <.0001 |
| Valve replacement/repair (n = 524) | 1,054.40 ± 347.49 | 1,430.35 ±<br>415.40 | <.0001 |
| Transplant (n = 19) | 1,083.11 ± 274.38 | 1,401.84 ±<br>272.50 | <.0001 |
| LVAD (n = 14) | 882.71 ± 416.18 | 1,204.64 ±<br>555.08 | <.01 |
| PAD (n = 42) | 863.86 ± 316.18 | 1,174.52 ±<br>427.52 | <.0001 |
| TAVR (n = 320) | 1,002.20 ± 362.23 | 1,287.61 ±<br>457.01 | <.0001 |
| Other (n = 113) | 1,096.50 ± 375.49 | 1,380.83 ±<br>401.44 | <.0001 |
| Diabetic (n = 1187) | 1,017.15 ± 358.33 | 1,343.35 ±<br>407.51 | <.0001 |

Abbreviations: CABG, coronary artery bypass graft; LVAD, left ventricular assist device; MI, myocardial infarction; PAD, peripheral artery disease; PCI, percutaneous coronary intervention; TAVR, transthoracic aortic valve replacement.

Similarly to 6MWT data, there was significant improvement of Dartmouth COOP scores across almost all referring diagnoses, except for those with LVAD diagnoses. Results can be visualized in Table 5. There was no difference in diabetic vs. non-diabetic in 6MWT scores (p = 0.08).

**Table 5.** Dartmouth COOP scores Pre and Post 12-63 Sessions of Cardiac Rehabilitation for the Referring Diagnosis and Diabetes.

| Referring Diagnosis | Pre | Post | P Value |
| --- | --- | --- | --- |
| MI (n = 264) | 21.44 ± 5.51 | 17.82 ± 5.26 | <.0001 |
| MI/CABG (n = 161) | 22.53 ± 5.44 | 16.50 ± 4.63 | <.0001 |
| MI/PCI (n = 757) | 21.16 ± 5.85 | 17.11 ± 4.93 | <.0001 |
| CABG (n = 830) | 22.21 ± 5.15 | 16.42 ± 4.42 | <.0001 |
| PCI (n= 997) | 21.59 ± 5.43 | 17.73 ± 4.94 | <.0001 |
| Angina (n= 118) | 23.35 ± 5.52 | 19.14 ± 4.95 | <.0001 |
| Heart Failure (n = 620) | 22.48 ± 5.61 | 18.67 ± 5.33 | <.0001 |
| Systolic heart failure (n = 352) | 22.58 ± 5.74 | 18.73 ± 5.62 | <.0001 |
| Diastolic/right heart failure (n = 183) | 22.22 ± 5.66 | 18.62 ± 4.94 | <.0001 |
| Valve replacement/repair (n = 524) | 22.34 ± 5.01 | 16.68 ± 4.24 | <.0001 |
| Transplant (n = 19) | 20.42 ± 4.88 | 15.95 ± 3.36 | <.0001 |
| LVAD (n = 14) | 21.50 ± 5.40 | 20.00 ± 5.16 | .17 |
| PAD (n = 42) | 22.67 ± 5.20 | 21.02 ± 4.81 | <.05 |
| TAVR (n = 320) | 20.82 ± 5.22 | 18.08 ± 4.89 | <.0001 |
| Other (n = 113) | 21.30 ± 5.24 | 17.86 ± 5.10 | <.0001 |
| Diabetic (n = 1187) | 22.59 ± 5.42 | 18.28 ± 5.09 | <.0001 |

## Discussion

This study demonstrated significant improvements in functional capacity and quality of life across patients undergoing phase II cardiac rehabilitation, as evidenced by increases in the six-minute walk test (6MWT) distance and reductions in Dartmouth COOP scores. The findings highlight the efficacy of CR programs in enhancing outcomes regardless of sex, race/ethnicity, or referring diagnoses.

### NUMBER OF PHASE II CR VISITS

The analysis of CR visits revealed a dose-response relationship, with greater improvements observed in patients completing more sessions. Significant gains in 6MWT were seen across all session tiers (12-23, 24-25, and 36+), with the most notable increases between the lowest and intermediate tiers. However, the diminishing marginal returns beyond 24-35 visits suggest a potential plateau effect, raising questions about the optimal number of CR sessions. These findings are consistent with prior studies and emphasize the importance of sustained engagement in CR to achieve maximal benefits^10–13^.

### SEX DIFFERENCES

Male participants constituted the majority (70.8%) of the cohort and exhibited higher pre- and post-CR 6MWT values compared to females. However, the relative improvements were significant for both sexes, suggesting CR benefits are not sex dependent. The lower baseline functional capacity in females aligns with prior research indicating sex-specific differences in physical performance and cardiovascular disease progression^14–16^. This underlines the importance of tailored interventions to address such disparities.

### RACIAL AND ETHNIC VARIABILITY

Participants were predominantly White (91.8%), with limited representation of minority groups, particularly Black (1.2%), American Indian (1.3%), and Asian participants (0.7%). While significant 6MWT improvements were observed across all racial groups, Black participants exhibited a statistically distinct percent change in 6MWT scores compared to others. These differences, coupled with lower baseline values for Black participants, may reflect underlying disparities in healthcare access or socioeconomic status^17,18^. Ethnic disparities were less pronounced, as improvements for Hispanic participants mirrored the overall cohort.

### IMPACT OF REFERRING DIAGNOSES

Patients with coronary artery bypass graft (CABG), percutaneous coronary intervention (PCI), or myocardial infarction (MI) represented the largest subgroups and showed substantial gains in functional capacity and quality of life. Conversely, patients with complex conditions such as left ventricular assist device (LVAD)^19^ placements or peripheral artery disease (PAD) exhibited lower baseline 6MWT scores and less pronounced improvements. While these gains remained statistically significant, they highlight the challenges faced by individuals with advanced cardiac pathology.

### STUDY LIMITATIONS

Several limitations warrant consideration. First, the dataset’s racial homogeneity limits generalizability to more diverse populations. Efforts to increase diversity should focus on recruiting more submitting practices from geographically and demographically varied regions. Additionally, future research should investigate potential barriers to CR participation within communities of color, including socioeconomic challenges and disparities in insurance coverage. Although most participants in this study had medical insurance, the data did not allow differentiation between private insurance, Medicare, and Medicaid, which could provide further insights into access and outcomes.

Additionally, the observational design of this study limits the ability to establish causal relationships between CR and the observed improvements in functional capacity and quality of life. While the results strongly suggest a positive impact of CR, a randomized controlled trial would provide more robust evidence by controlling potential confounding variables such as baseline health status, socioeconomic factors, and healthcare access. This would also allow for better assessment of whether observed improvements are directly attributable to CR or influenced by external factors, such as concurrent medical therapies or lifestyle changes.

Finally, the analysis of subgroup performance based on the number of CR sessions revealed diminishing returns beyond 24-35 visits, suggesting a potential plateau in functional capacity improvement. However, the precise threshold for achieving optimal benefits remains unclear. Individual variability, such as comorbid conditions, baseline functional status, and adherence to CR protocols, may influence the required number of sessions for maximal improvement. Future studies should aim to identify personalized session targets and explore whether certain subgroups benefit from extended CR programs compared to others.

Additionally, the observational design restricts causal inferences about the observed improvements. A randomized controlled trial would better delineate the direct effects of CR. Third, while the Dartmouth COOP is a validated tool, its subjective nature may introduce bias. Additionally, subgroup analyses by the number of CR sessions suggest diminishing returns beyond 24-35 visits, yet the precise threshold for optimal benefit remains unclear.

## Conclusion

This study highlights the substantial benefits of phase II cardiac rehabilitation in improving functional capacity and quality of life among patients with cardiovascular disease. Significant gains were observed across all patient subgroups, with functional capacity improvements influenced by the number of CR sessions completed. While a dose-response relationship was evident, diminishing returns were noted beyond 24-35 visits, suggesting the need for further investigation into individualized session targets.

Demographic factors, including sex and race/ethnicity, also played a role in observed outcomes, with disparities pointing to the need for tailored strategies to enhance access and effectiveness for underrepresented populations. Additionally, referring diagnoses impacted functional capacity improvements, with patients presenting with advanced or complex conditions showing smaller, albeit still significant, gains.

Despite the study’s limitations, these findings reinforce the critical role of exercise-based CR in managing cardiovascular disease. Efforts to expand access, address barriers in underserved communities, and explore individualized approaches to CR dosing are essential for maximizing the program’s impact on diverse patient populations.

## Data Availability

All data produced in the present study are available upon reasonable request to the authors.

## Author Declaration

The authors wish to confirm that there are no known conflicts of interest associated with this publication and there has been no significant financial support for this work that could have influenced its outcome.

## Notes

### Competing Interest Statement

The authors have declared no competing interest.

### Funding Statement

This study did not receive any funding.

### Author Declarations

Datasets used were de-identified prior to use in our study.

